# Language Uncovers Visuospatial Dysfunction in Posterior Cortical Atrophy: A Natural Language Processing Approach

**DOI:** 10.1101/2023.11.21.23298864

**Authors:** Neguine Rezaii, Daisy Hochberg, Megan Quimby, Bonnie Wong, Scott McGinnis, Bradford C Dickerson, Deepti Putcha

## Abstract

**Introduction:** Posterior Cortical Atrophy (PCA) is a syndrome characterized by a progressive decline in higher-order visuospatial processing, leading to symptoms such as space perception deficit, simultanagnosia, and object perception impairment. While PCA is primarily known for its impact on visuospatial abilities, recent studies have documented language abnormalities in PCA patients. This study aims to delineate the nature and origin of language impairments in PCA, hypothesizing that language deficits reflect the visuospatial processing impairments of the disease.

**Methods:** We compared the language samples of 25 patients with PCA with age-matched cognitively normal (CN) individuals across two distinct tasks: a visually-dependent picture description and a visually-independent job description task. We extracted word frequency, word utterance latency, and spatial relational words for this comparison. We then conducted an in-depth analysis of the language used in the picture description task to identify specific linguistic indicators that reflect the visuospatial processing deficits of PCA.

**Results:** Patients with PCA showed significant language deficits in the visually-dependent task, characterized by higher word frequency, prolonged utterance latency, and fewer spatial relational words, but not in the visually-independent task. An in-depth analysis of the picture description task further showed that PCA patients struggled to identify certain visual elements as well as the overall theme of the picture. A predictive model based on these language features distinguished PCA patients from CN individuals with high classification accuracy.

**Discussion:** The findings indicate that language is a sensitive behavioral construct to detect visuospatial processing abnormalities of PCA. These insights offer theoretical and clinical avenues for understanding and managing PCA, underscoring language as a crucial marker for the visuospatial deficits of this atypical variant of Alzheimer’s disease.

## Introduction

Posterior cortical atrophy (PCA) is a clinico-radiological syndrome characterized by a progressive decline in higher-order visuospatial processing with relative preservation in other cognitive domains at initial presentation (1–3). Common visuospatial symptoms of the syndrome include impaired object and space perception, simultanagnosia, environmental agnosia, and visual field defects (2). From the neuroimaging perspective, the syndrome is associated with atrophy, hypometabolism, and usually tau deposition in posterior parietal, occipital, and temporo-occipital cortices (4,5). As the majority of PCA cases are due to underlying Alzheimer’s pathology, PCA is also referred to as the visual variant of Alzheimer’s disease (AD) (6,7). While the diagnostic criteria for PCA indicate preserved functions in cognitive domains outside of visuospatial processing at symptom onset, a growing literature has documented language abnormalities in PCA emerging early in the course of the illness. Specifically, impaired category fluency and confrontation naming have been documented on formal neuropsychological assessments (8–11). Language abnormalities are also evident during spontaneous speech, such as using higher frequency words and slowed speech rate (number of words per minute) (12). These emerging observations suggest that there is still much to be understood about the nature and origin of language impairments in PCA.

The specific types of language abnormalities observed in PCA may be related to the network dysfunction that supports lexicosemantic retrieval, as has been previously postulated across the phenotypic spectrum of AD (10). Another possible explanation is that the language abnormalities observed in PCA may stem from the visuospatial impairments central to the syndrome, rather than representing a primary language deficit. A large body of research supports close relationships between the visual processing of objects and the amodal semantic processing required for retrieving the names of those objects (13–15). Recently, it has been shown that anterior to each region that is selective for the visual processing of a given category in the visual cortex, there is a corresponding area selective to its linguistic processing (16). This anatomical and functional configuration suggests that the anterior border of the visual cortex acts as a convergence zone where information from the unimodal visual system enters the amodal linguistic systems involved in linguistic retrieval. Therefore, the successful production of a word that has visual attributes requires intact visual processing, essential for providing the information needed to retrieve its corresponding linguistic representation (i.e., its name). Therefore, deficits in visual processing would theoretically impede the production of words with visual attributes. If the pathophysiology of language abnormalities in PCA involves disrupted visual processing, then it stands to reason that tasks heavily dependent on visual processing will exhibit significant language impairments. Conversely, tasks with minimal reliance on visual input should result in relatively intact language performance.

In the current study, we sought to test this hypothesis by contrasting the language used in two different speech samples as PCA participants described the Picnic scene from the Western Aphasia Battery (17) (visually dependent) and their prior jobs (visually independent). In each speech sample, we measured word frequency, word utterance latency, and the use of spatial relational words. For the picture description task, we hypothesized that PCA patients would use higher frequency words (e.g., replacing specific names of pictured items with superordinate words potentially including “thing”), have increased word utterance latency due to object recognition difficulty, and use fewer spatial relational words such as “into” or “underneath” compared to healthy individuals. For the non-visually dependent job description task, we expected these linguistic features to be comparable between PCA patients and healthy individuals.

Building on the hypothesis that speech patterns in visually dependent tasks reflect visuospatial processing deficits, we next sought to identify linguistic markers of these challenges. Specifically, we investigated which elements in the picnic scene presented particular retrieval difficulties and whether PCA patients could intuitively grasp and articulate the overall theme of the scene, such as using the word “picnic”. Due to the difficulty with visually integrating a scene (simultanagnosia) (9,18,19), we hypothesized that PCA patients were less likely to verbalize the term “picnic” compared to healthy individuals. Lastly, to address the clinical significance of this work, we used the language features derived from the picture description task to develop a classifier aimed at distinguishing PCA patients from healthy individuals and hypothesized a high degree of classification accuracy.

## Methods

### Participants

#### PCA patients

Twenty-five patients diagnosed with PCA were recruited from the Massachusetts General Hospital (MGH) Frontotemporal Disorders Unit PCA program for this study (20). All but one was confirmed amyloid positive (A+) and tau positive (T+) by either CSF analysis or amyloid and tau PET. The remaining participant’s biomarker status is unknown due to a failed lumbar puncture. Each patient had posterior cortical atrophy and/or hypometabolism, consistent with the typical neurodegeneration (N+) of PCA. See Table 1 for demographic and clinical data. All participants received a standard clinical evaluation comprising a structured history obtained from both participant and informant, comprehensive neurological and psychiatric history, as well as neuropsychological assessment. See Table S1 for neuropsychological profiles of the PCA cohort included in this study. The clinical formulation was performed through a consensus conference by our multidisciplinary team of neurologists, psychiatrists, neuropsychologists, and speech and language pathologists, with each patient classified based on all available clinical information as having a 3-step diagnostic formulation of mild cognitive impairment or dementia (Cognitive Functional Status), a specific Cognitive-Behavioral Syndrome, and a likely etiologic neuropathologic diagnosis (21). Patients underwent neuroimaging sessions involving structural MRI, FTP PET, and amyloid (PiB or FBB) PET scans of CSF analysis for Aβ and tau. Aβ positivity was determined by a combination of visual read and mean amyloid PET signal extracted from a cortical composite region of interest according to previously published procedures (22–24). Determination of tau and neurodegeneration positivity was conducted by visual read using internal methods similar to published work (25–27). This work was carried out according to The Code of Ethics of the World Medical Association (Declaration of Helsinki) for experiments involving humans. All participants and their caregivers provided informed consent in accordance with the protocol approved by the Mass General Brigham Human Research Committee Institutional Review Board in Boston, Massachusetts. A speech sample for the picture description task was acquired from all twenty-five PCA participants. Twenty-three PCA participants also took part in the job description task.

**Table 1.** Clinical characteristics of the Aß+ Posterior Cortical Atrophy (PCA) and Cognitively Normal (CN) groups. Means and Standard Deviations are reported for continuous variables. MoCA = Montreal Cognitive Assessment. CDR = Clinical Dementia Rating Scale. SOB = Sum of Box scores.

| Demographics | PCA (N = 25) | CN1 (N = 29) | CN2 (N = 26) |
| --- | --- | --- | --- |
| Age (years) | 68.4 ± 7.8 | 65.3 ± 8.4 | 61.8 ± 7.8 |
| Sex (M/F) | 13/12 | 13/16 | 9/17 |
| Education (years) | 17.2 ± 2.1 | 15.7 ± 1.0 | 14.8 ± 1.8 |
| Handedness<br>(R/L/Ambidextrous) | 23/0/2 | 20/8/1 | 20/3/3/ |
| MoCA | 14.6 ± 7.8 | -- |  |
| CDR | CDR 0 (N=1)<br>CDR 0.5 (N=11)<br>CDR 1 (N=10)<br>CDR 2 (N=3) | -- |  |
| CDR-SOB | 4.5 ± 3.1 | -- |  |

#### Cognitively normal (CN) individuals

Twenty-nine CN participants (CN1) were enrolled through the Speech and Feeding Disorders Laboratory at the MGH Institute of Health Professions to participate in the picture description task. These participants passed a cognitive screen, were native English speakers, and had no history of neurologic injury or developmental speech/language disorders. Twenty-six CN participants (CN2) were additionally recruited through Amazon’s Mechanical Turk (MTurk) to describe their jobs. MTurk participants filled out the short and validated version of the 12-item Everyday Cognition questionnaire, a questionnaire designed to detect cognitive and functional decline (28). Only language samples from participants who were native English speakers with no self-reported history of brain injury or speech/language disorder, either developmental or acquired, were included in the analyses.

### Speech samples and data analysis

Speech samples were collected under two conditions. For the visually dependent task, participants described the Western Aphasia Battery—Revised (WAB-R) (17) Picnic Scene with the instruction to use full sentences. For the visually independent task, participants were asked to describe what they did for work. There were no time limits applied to either task. Autotranscription was done using Google Cloud Speech-to-Text API for audio transcription (29), and manually verified by a research staff blinded to the diagnosis.

#### Speech sample analysis

All feature extraction was performed automatically using Quantitext, a speech and text analysis toolbox we developed in the Frontotemporal Disorders Unit of MGH, to produce a set of quantitative speech and language metrics. The goal of developing this package is to increase the precision and objectivity of language assessments along the goals described previously (30). The toolbox incorporates several natural language processing toolkits and software such as Stanza (31) as well as text analysis libraries in R. Quantitext receives audio or transcribed language samples as input and generates a number of metrics such as word frequency, syntax frequency (32), content units (33), total units, efficiency of words (34), and part-of-speech tags.

#### Word frequency

To measure word frequency, we used the Switchboard corpus (35), which consists of spontaneous telephone conversations averaging six minutes in length spoken by more than 500 speakers of both sexes from a variety of dialects of American English. We use this corpus to estimate word frequency in spoken English, independently of the patient and control sample. The corpus contains 2,345,269 words. Here, word frequency denotes the log frequency of content words (comprised of nouns, verbs, adjectives, and adverbs).

#### Word utterance latency and articulation rate

Our analysis employed the Google Cloud Speech-to-Text API to ascertain word timestamps, pinpointing the onset and offset for each spoken word within the audio recordings. Speech rate—often quantified as the number of words spoken per minute—can vary based on factors such as word utterance latency and the individual articulation rate of each word. To ensure a more granular and accurate interpretation of the underlying phenomena, we sidestepped aggregated metrics like speech rate, focusing instead on separately evaluating its constituent components. Word utterance latency is defined as the time interval preceding the articulation of a word. This method was applied on all except for the very first word in each sample, as the time to start the description task depends on multiple factors. Articulation rate measures the number of syllables per second (36).

#### Spatial relational words

Relational words are automatically tagged by Stanza as “case”. For most words, the relational words are spatial, for example, the word “under” in the sentence “I found the gem under my bed”. In our analysis, we divided the number of relational words by the total words.

#### Content units

To determine which items within the picture posed greater challenges for PCA compared to CN participants, we coded the visual items using content units. Content units are words with correct information units that are intelligible in context and accurate about the picture or topic. Words do not have to be used in a grammatically correct manner to be counted as content units (37). Each content unit is only counted once, regardless of how many times it is mentioned in a sample. The morphological variants were grouped within one single content unit. For example, the nouns ‘girl’ and ‘daughter’ are considered the same content unit. Therefore, if one participant used both words (girl and daughter), they would only be counted as one content unit. To specify content units, Quantitext first generates a Python dictionary using a predefined set of words as previously described and then uses this dictionary to automatically identify all content units in new texts it receives. Previously, we showed that the program has an accuracy of 99.7% in identifying content units (33).

#### Statistical analysis

We used Welch Two Sample t-tests to compare the language features across the two groups. We performed a pairwise Pearson correlation analysis on our dataset to investigate the relationships between the likelihood of reporting each content unit and the group designation (with PCA dummy coded as 1 and healthy controls as 0). For classification, we used a binary logistic regression model. We employed a leave-one-out cross-validation (LOOCV) approach on our dataset to validate the model’s performance. In each iteration of the LOOCV, a single observation was set aside as the test data, and the remaining observations were used to train the model. Alpha was set at 0.05.

## Results

### Language abnormalities in PCA are observed during picture description but not job description

We first compared the speech samples of PCA participants describing the WAB Picnic Scene to the CN1 group to determine language abnormalities in this task. We also compared the speech sample from the job description task between the PCA and CN2 groups. We used Welch Two Sample t-tests to compare the means of the following features across the two groups (Figure 1).

**Figure 1.**
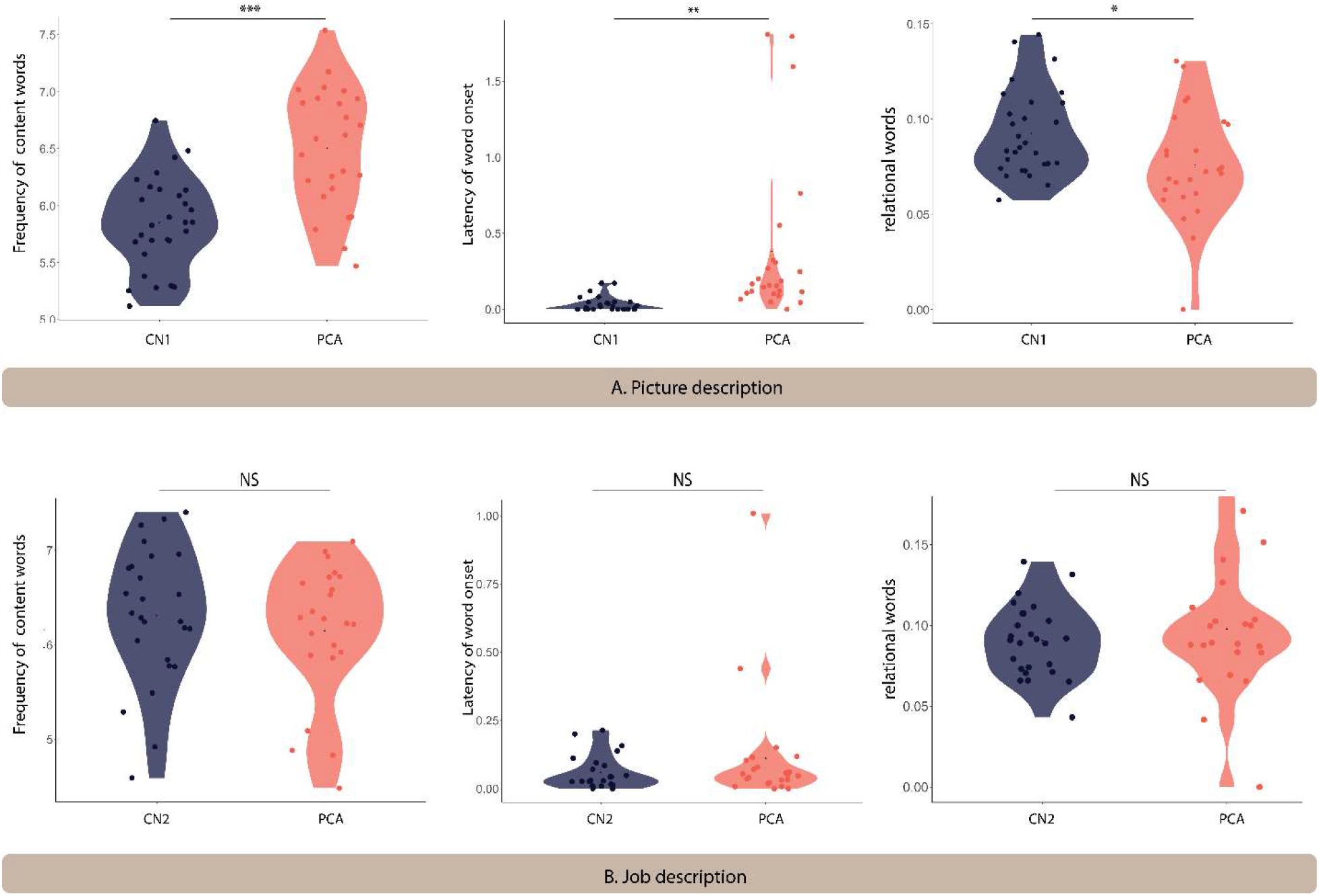
Language differs between PCA and healthy controls on the picture description task but not the job description task. Violin plots comparing the language features extracted from the picnic scene **(A)** and job **(B)** description tasks across healthy controls and PCA patients. *** denotes p < 0.001, ** indicates 0.001 < p < 0.01, and * shows 0.01 < p < 0.05. NS indicates not significantly different.

#### Picture description task

Patients with PCA used higher frequency words (i.e., more commonly used words) (mean = 6.50 ± 0.53) compared to healthy controls (mean = 5.85 ± 0.40) (t(44.28) = -5.02, p < 0.001). The time latency to the onset of words was longer for patients with PCA (mean = 0.38 ± 0.54) compared to healthy controls (mean = 0.03 ± 0.05) (t(24.34) = -3.20, p = 0.004). We found that patients with PCA used fewer spatial relational terms (mean = 0.08 ± 0.03) compared to healthy controls (mean = 0.09 ± 0.02) (t(45.55) = 2.35, p = 0.023).

There was a trend toward a slower articulation rate in PCA patients (mean = 2.92 ± 0.60) compared to healthy controls (mean = 3.14 ± 0.35) (t(33.88) = 1.616, p = 0.115), suggesting that slower speech rate may be primarily due to an increased word utterance latency rather than articulation rate.

#### Job description task

The frequency of content words in PCA patients (mean = 6.15 ± 0.72) was not different from healthy controls (mean = 6.31 ± 0.72) (t(46.27) = 0.79, p = 0.43). Similarly, there was no significant difference in word utterance latency between PCA patients (mean = 0.11 ± 0.22) and healthy controls (mean = 0.06 ± 0.06) (t(25.24) = -1.11, p = 0.28). No statistical difference was found in the use of relational words between PCA patients (mean = 0.10 ± 0.04)) and healthy individuals (mean = 0.09 ± 0.02) (t(33.36) = - 0.74, p = 0.47). There was no difference in articulation rate in PCA patients (mean = 2.93 ± 0.60) compared to healthy controls (mean = 3.11 ± 0.35) (t(39.76) = 1.12, p = 0.269).

### Specific language indicators of visuospatial processing deficits of PCA can be extracted from the picture description task

We next probed the samples obtained from the picture description task to extract the specific language features that reflect visuospatial impairment in PCA compared to healthy controls. First, we determined the likelihood of mentioning each content unit by each diagnostic group. The picture consists of 32 content units, as shown in Figure 2. We performed a pairwise Pearson correlation analysis on our dataset to investigate the relationships between the likelihood of reporting each content unit and the group designation. As shown in Figure 2, we did not observe a uniform reduction in the likelihood of mentioning each content unit in PCA. Instead, certain content units had a much lower chance of being verbalized. Of all content units, “fisherman”, a small, central feature of the WAB Picnic scene, was the least likely to be mentioned by a patient with PCA compared with healthy controls (r = -0.85, p < 0.001). A few content units had a numerically higher, though not statistically significant, likelihood of being mentioned by patients with PCA compared to healthy individuals, such as “clouds” (r = 0.18, p = 0.186). Figure 3 is the artistic rending we developed to show the rate at which patients with PCA mention each content unit. We then compared the total number of content units retrieved across the two groups. Overall, PCA patients retrieved fewer content units (mean = 7.20 ± 5.63) compared to healthy individuals (mean = 16.28 ± 4.41) (t(45.21) = 6.52, p < 0.001). Lastly, patients with PCA had a lower likelihood of reporting the overall theme of the picture (i.e., mentioning the word “picnic”) (mean = 0.16 ± 0.37), than healthy individuals did (mean = 0.90 ± 0.31) (t(46.77), p < 0.001).

**Figure 2.**
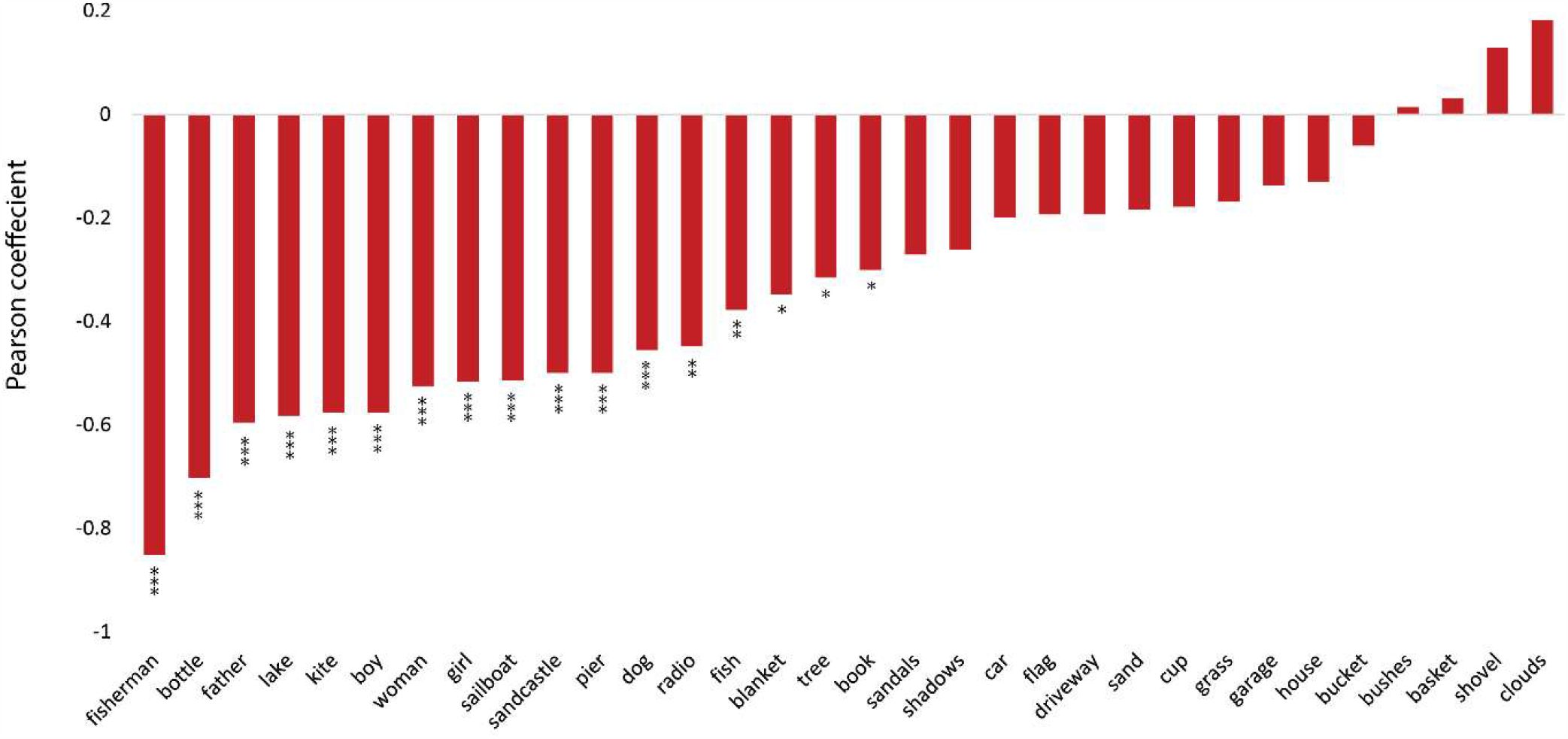
PCA participants and healthy individuals described different content units on the picture description task. The Pearson correlation coefficients between the likelihood of reporting each content unit in the picnic scene and the designated group. Negative values indicate that PCA patients have a lower chance of mentioning the content unit compared to healthy individuals. *** denotes p < 0.001, ** indicates 0.001 < p < 0.01, and * shows 0.01 < p < 0.05. Bars without an asterisk are not significantly different.

**Figure 3.**
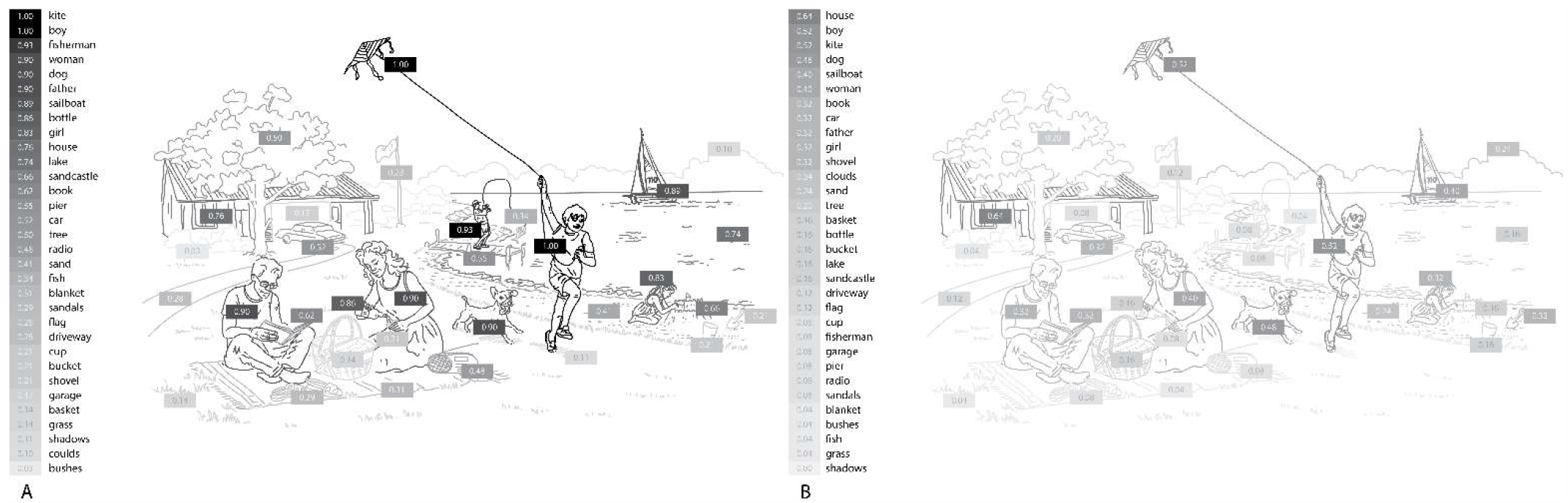
The likelihood of mentioning each content unit of the WAB Picnic Scene by PCA patients and healthy individuals. The shading intensity of each item corresponds to its verbalization probability by participants, with darker elements indicating a higher likelihood of being mentioned by PCA patients and healthy individuals.

### Diagnostic Classification

We used binary logistic regression to classify PCA and healthy individuals. Our predictor variables consisted of word frequency, word utterance latency, relational words, the total number of content units, and the probability of mentioning “picnic”. As the sixth variable, we included the probability of mentioning “fisherman” because this content unit had the highest correlation with the group designation, likely due to its visuospatial processing demands. The average accuracy of the model was 98.15% after leave-one-out cross-validation. The average precision across all iterations was found to be 0.96, which means that, on average, 96% of the predicted positive cases were actual positive cases. Moreover, the model demonstrated an average recall of 1, indicating that it successfully identified all the positive cases from the test data in each iteration. We also evaluated the performance of the model using a Receiver Operating Characteristic (ROC) curve. The Area Under the Curve (AUC) was 1, indicating the perfect discrimination ability of the model (Figure 4).

**Figure 4.**
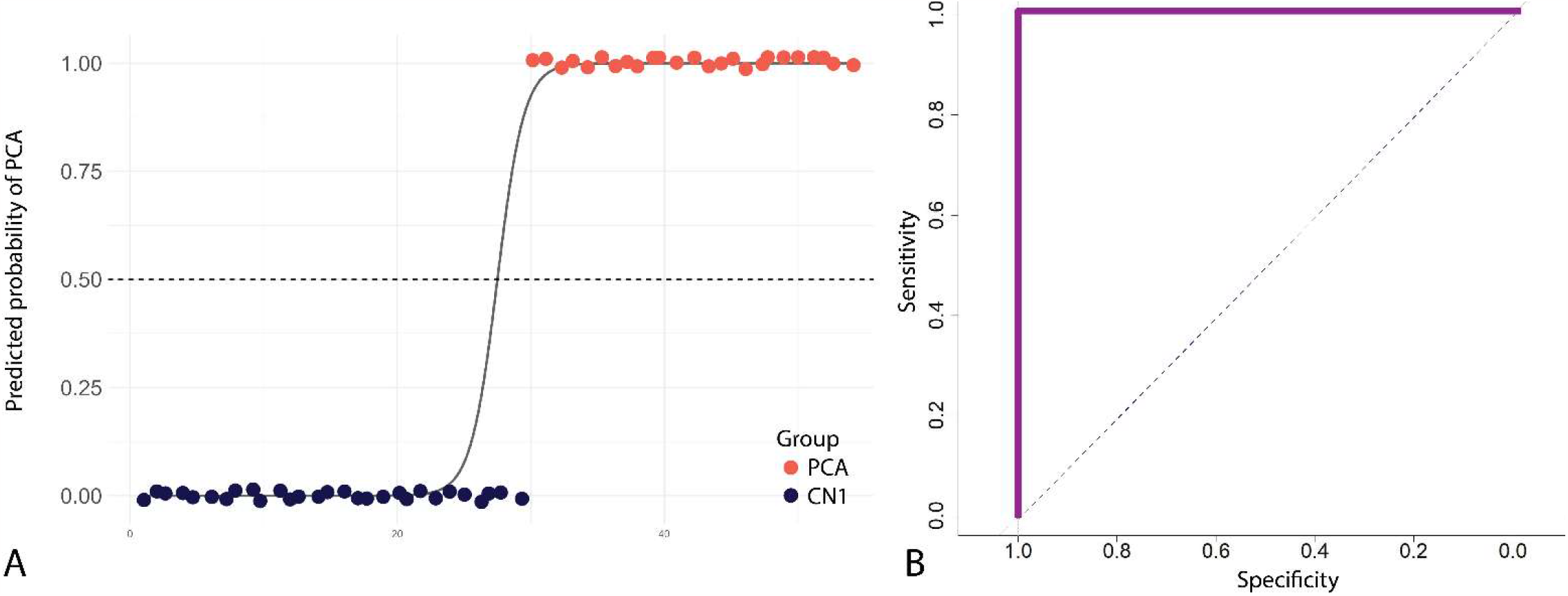
Diagnostic performance of a model that distinguishes PCA patients from healthy individuals using linguistic features from the picture description task. **(A)** The scatter plot shows the predicted probabilities of classifying an individual as having PCA using selected linguistic features. Each point represents an individual participant, with colors indicating the group. The sigmoid curve illustrates the general trend of predicted probabilities. The dashed black line at the predicted probability of 0.5 serves as a decision threshold to classify between PCA patients and healthy individuals. Points are jittered vertically for better visualization. **(B)** ROC curve illustrating the performance of the logistic regression model in discriminating between PCA patients and healthy individuals, with an AUC value of 1 indicating the model’s overall perfect accuracy.

## Discussion

Using computational linguistic methods, our study illuminated a distinction in PCA patients between compromised language abilities in visually-dependent tasks and preserved language skills in visually-independent contexts. At the theoretical level, we used this method to show that at least some of the language abnormalities increasingly being identified in PCA are byproducts of visuospatial deficits characteristic of this atypical AD syndrome. On the visually-independent job description task, the characteristics of language production we measured here were not impaired in patients with PCA. Translating our observations to clinical practice, we showed that computational linguistic analysis of a simple picture description task robustly classified nearly all PCA patients as distinct from healthy controls, supporting its value in clinical diagnostic evaluation.

Our work is consistent with studies showing rich connections between networks representing information directly received from senses and information conveyed through spoken language (16,38,39). Unimodal sensory information and abstract language information are combined at multiple points across the cortex, such as inferior parietal lobule (comprising the angular and supramarginal gyri) and large swaths of posterolateral temporal cortex (40,41), many of which can be affected in PCA. Therefore, language abnormalities in PCA may arise for at least two potential reasons. First, the neurodegeneration of PCA may extend beyond visuospatial areas to encompass regions involved in abstract language processing. In support of this hypothesis, evidence suggests that the brain regions affected in PCA overlap with those critical for word retrieval (42–46). Alternatively, language anomalies may arise as a consequence of visuospatial deficits hindering the transfer of necessary sensory information for amodal language processing.

While these two possibilities are not mutually exclusive, our results suggest that language impairments might be largely secondary to visuospatial dysfunction. In our analysis comparing a variety of speech and language properties of the narratives produced when PCA patients describe a complex visual scene versus a recounting of their primary occupation from memory, we observed speech and language impairments in only the visually dependent picture description task. We believe the increased word frequency and word utterance latency in the picture description task reflect visual deficits in object recognition. Similarly, the reduced use of spatial relational words may reflect the patients’ difficulty processing spatial relations between elements of the picnic scene. The absence of abnormalities in word frequency, word utterance latency, and relational words in the job description task provides evidence that these language impairments do not stem from an intrinsic deficit in the language system in PCA. Since most daily communication is a blend of visuospatial cognition, episodic memory, and other cognitive domains, we anticipate that an analysis of everyday speech would reveal linguistic deficiencies proportionate to the visuospatial load of its content. This expectation aligns with prior research reporting linguistic anomalies in participants recounting their recent holiday, an account that naturally encompasses the visuospatial processing of a recent event, such as where they went and what they saw (12). Another consideration in interpreting these task differences is that the picture description task required the use of specific linguistic elements representing the specific visual stimulus. It is also possible that when given fewer constraints in the job description task, individuals had the freedom to choose a potentially more familiar and more easily accessible language. Our explanation of the underlying language abnormalities in PCA is consistent with findings that showed a striking discrepancy between visual and verbal comprehension tasks in this population (47). Our conclusion is also synergistic with results reporting very mild impairment in semantic memory in PCA, indicating that the apparent semantic impairment in these conditions may be secondary to visual impairment (47).

Based on the observation that language is a sensitive indicator of visuospatial impairments of PCA, we performed an in-depth content unit analysis of language elicited through the picture description task. First, we measured the probability of verbalizing each content unit of the picture. The most distinguishing feature between PCA patients and healthy individuals was the probability of mentioning the “fisherman”. While 93% of healthy participants mentioned this content unit, only 8% of PCA patients did so. This discrepancy could be attributed to the smaller size of this element in the picture. In addition, multiple elements are superimposed in the location of this content unit. Numerically, though not significantly, certain items, such as “clouds,” had a higher likelihood of being mentioned by PCA patients than healthy individuals. This type of analysis provides a naturalistic way of identifying the visuospatial elements that are particularly challenging for PCA patients and could help clinicians devise rehabilitative strategies to alleviate these challenges. Moreover, we observed that PCA patients often missed describing the overarching theme of the image (“picnic”), even when they identified certain components related to it (“basket”). We believe this finding represents the effects of simultagnosia, which prevents many PCA patients from grasping the integrated theme of a composite visual entity.

Finally, when specific quantitative language metrics were employed to differentiate PCA patients from healthy individuals, our predictive model achieved a high level of performance, as evidenced by an AUC of 1 and an accuracy rate of 98.15%. Automating this linguistic evaluation from an easily acquired speech sample would facilitate the integration of measures like this into digital healthcare infrastructure, which a wide array of healthcare providers could potentially use once trained. This development has potential implications for improving early diagnosis of PCA as well as for monitoring response to disease-modifying, rehabilitative, or other therapies in this underserved atypical variant of AD.

## Data Availability

All data produced in the present study are available upon reasonable request to the authors

## Author contributions

NR, DP contributed to the conception, design of the study, and data analyses. NR, DP, and BCD drafted the manuscript. NR, DP, DH, MQ, BW, SM, and BCD participated in the data collection and editing of the manuscript.

## Data availability

Language data and codes will be available upon request to Dr. Bradford C Dickerson.

